# Biomarker Identification in Pancreatic Cancer Through Concordant Differential Expression and Interpretable Machine Learning Analyses

**DOI:** 10.64898/2026.02.13.26346263

**Authors:** Sonia Maciá-Escalante, Rubén López-Aladid, Rebeca Muñoz-Tovar, Manuel López-Herrero, Ana Navarro-Sellés, Leonor Garmendia, Carolina Puerto, Mariana Fossati-Vázquez, Pablo Parente-Muñoz

## Abstract

**Background:** Pancreatic ductal adenocarcinoma is one of the most aggressive and lethal malignancies of the gastrointestinal tract. The poor prognosis is largely attributed to late-stage diagnosis, pronounced tumor heterogeneity, and limited therapeutic efficacy. These challenges underscore the urgent need for the identification of robust molecular biomarkers and novel therapeutic targets.

**Methods:** Gene expression data from a total of 146 pancreatic tissue samples, comprising 72 normal and 74 tumor specimens obtained from the Pan-Cancer Atlas(TCGA) were analyzed. Differential gene expression analysis was conducted using the DESeq2 package, followed by functional enrichment analysis based on GO and KEGG. A classification model was developed using the XGBoost algorithm and evaluated through 500 bootstrapping iterations and 5-fold cross-validation to ensure robustness and generalizability. Model interpretability was assessed using SHAP (SHapley Additive exPlanations) values to identify genes with the highest predictive contribution.

**Results:** A comprehensive transcriptomic analysis revealed significant dysregulation of multiple genes between normal and tumor pancreatic tissues. Genes such as GJB3, S100A2, MSLN, and SLC2A1 were notably overexpressed, whereas DEFA6, APOB, and RBP2 exhibited marked downregulation, indicative of impaired exocrine function and aberrant epithelial reprogramming. The XGBoost classification model achieved an average area under the curve (AUC) of 0.9868 and an overall accuracy of 98.6%. SHAP (SHapley Additive exPlanations) analysis identified GJB3, LINC02086, and TSPAN1 as key predictive features. Six genes were concurrently identified as differentially expressed and highly influential within the model, supporting their potential utility as robust biomarkers for pancreatic tumor characterization.

**Conclusions:** Pancreatic ductal adenocarcinoma is marked by extensive transcriptomic reprogramming. The integration of differential gene expression analysis with interpretable machine learning enabled the identification of a molecular signature with potential diagnostic and therapeutic relevance.

## Introduction

Pancreatic ductal adenocarcinoma (PDAC) remains one of the most lethal malignancies worldwide, accounting for more than 90% of all pancreatic cancer cases and representing the seventh leading cause of cancer-related death globally [1]. Its dismal five-year survival rate, which remains below 12%, is primarily attributed to the asymptomatic nature of early disease, late clinical presentation, and the intrinsic resistance of tumor cells to conventional therapies [2,3]. Current clinical guidelines from the National Comprehensive Cancer Network (NCCN), the European Society for Medical Oncology (ESMO), and the American Society of Clinical Oncology (ASCO) emphasize multimodal strategies combining surgical resection, systemic chemotherapy, and radiotherapy in selected cases [4–6]. However, only 15–20% of patients present with resectable disease at diagnosis, and recurrence rates remain exceedingly high even after curative-intent surgery and adjuvant chemotherapy with FOLFIRINOX or gemcitabine-based regimens [7]. Thus, there is an urgent need for reliable biomarkers that enable early detection, accurate prognostic stratification, and personalized therapeutic decision-making.

Recent advances in molecular oncology have illuminated key genetic alterations in PDAC, including mutations in KRAS, TP53, CDKN2A, and SMAD4, which drive tumorigenesis and resistance mechanisms [8]. Despite these insights, the translation of molecular discoveries into clinical applications has been limited, partly due to the profound inter- and intra-tumoral heterogeneity of PDAC [9]. Transcriptomic profiling through RNA sequencing (RNA-seq) has emerged as a powerful approach to comprehensively assess gene expression patterns and uncover novel molecular subtypes of PDAC [10]. Such analyses have revealed deregulation of pathways related to epithelial-mesenchymal transition, immune evasion, and metabolic reprogramming, providing a foundation for biomarker discovery and potential therapeutic targets [11]. Nevertheless, the high dimensionality and complexity of transcriptomic datasets necessitate advanced computational approaches capable of distinguishing true biological signals from noise.

Machine learning (ML) approaches have demonstrated remarkable utility in deciphering complex omics data, offering predictive frameworks that complement traditional biostatistical methods. In particular, algorithms such as extreme gradient boosting (XGBoost) have been successfully applied to classify tumor phenotypes and predict clinical outcomes with high accuracy [12]. To enhance interpretability, the integration of explainability frameworks—such as SHapley Additive exPlanations (SHAP)—has facilitated the identification of key molecular features driving model predictions, thereby increasing their biological and clinical relevance [13]. This intersection between ML and transcriptomics represents a transformative paradigm in precision oncology, with the potential to refine diagnostic criteria and guide targeted therapeutic interventions.

In this study, we performed a comprehensive transcriptomic analysis comparing normal and tumor pancreatic tissues to identify differentially expressed genes (DEGs) associated with PDAC. Functional enrichment analyses were conducted to delineate the biological pathways disrupted in tumorigenesis. Furthermore, we implemented an XGBoost classification model integrated with SHAP-based interpretability to prioritize genes with strong discriminative power and biological significance. Our results highlight GJB3 and LINC02086 as potential dual-significance biomarkers with both diagnostic and mechanistic relevance, warranting further experimental validation and evaluation in clinical cohorts.

## Methods

### Study design

We conducted a cross-sectional computational study integrating differential gene expression (DGE), pathway enrichment, supervised classification, and model explainability to discriminate pancreatic ductal adenocarcinoma (PDAC) from normal pancreas using bulk RNA-sequencing (RNA-seq).

### Data source and participants

Public, de-identified RNA-seq count matrices were obtained from the TCGA Pan-Cancer Atlas (pancreas). The analytic cohort comprised primary PDAC tumors and histologically normal pancreatic tissues (total n=146; 74 tumor, 72 normal). Inclusion criteria were unambiguous tumor/normal labels, availability of sequencing quality metadata, and ≥10 million mapped reads. Only primary tumors were considered. Samples failing prespecified quality thresholds (library outliers, marked 5′–3′ bias, or inadequate gene-body coverage) were excluded.

### RNA-seq processing and quality control

Gene-level Ensembl counts were processed following a prespecified pipeline. Library quality was assessed by library size distributions, duplication rates, 5′–3′ bias, and gene-body coverage. Lowly expressed features were filtered using a counts-per-million rule (CPM≥1 in ≥70% of samples in at least one class). rRNA and pseudogenes were removed. Ensembl identifiers were mapped to HGNC symbols using one-to-one mappings. For DGE, raw counts were normalized with DESeq2 median-of-ratios size factors. When center/batch labels were available, they were modeled as fixed effects in the design; when absent, surrogate variable analysis was considered. We did not apply ComBat to counts destined for differential testing.

### Differential gene expression

Differential expression was performed on raw gene-level counts using DESeq2. Wald tests were performed and P values were adjusted using the Benjamini–Hochberg false discovery rate (FDR). Significance was defined a priori as FDR<0.05 with absolute log2 fold change (|log2FC|) ≥2. We generated volcano and MA plots (apeglm shrinkage) and conducted principal component analysis on variance-stabilized data to visualize class separation and residual batch effects.

### Functional enrichment

Over-representation analysis (ORA) was performed on the expressed gene universe using clusterProfiler for Gene Ontology (GO Biological Process 2021) and KEGG 2021. Up- and down-regulated gene sets were analyzed separately with FDR control at q<0.05. To reduce term redundancy, GO results were pruned by semantic similarity (cutoff 0.7). Enrichment was summarized by dot plots and gene–term network diagrams.

### Machine-learning classification

An extreme gradient boosting (XGBoost) classifier was trained on variance-stabilized expression. All preprocessing was nested within each training split to prevent information leakage. Within each bootstrap replicate, expression values were quantile-transformed to approximately Gaussian margins and z-scaled; features were preselected by variability (interquartile-range percentile threshold) and highly collinear genes were pruned by greedy removal at |Pearson r|>0.95. Model evaluation used 500 stratified bootstraps with 75/25 train–test splits. Hyperparameters were tuned by five-fold inner cross-validation with early stopping to maximize the area under the receiver operating characteristic curve (AUROC). The primary outcome was AUROC on held-out tests; secondary outcomes were accuracy, precision, recall, F1 score, area under the precision–recall curve, and calibration (Brier score and reliability curves). We report mean (SD) and percentile-based 95% intervals across bootstraps. A parsimony analysis retrained a reduced model using only the top SHAP-ranked genes, with the number of features chosen by an elbow analysis of AUROC versus feature count.

### Model explainability

Model interpretability used TreeExplainer (interventional SHapley Additive exPlanations [SHAP]) applied to each bootstrap’s final model on the corresponding held-out test set. Global importance was summarized as mean absolute SHAP per gene, aggregated across test samples and across bootstraps. Directionality was assessed with SHAP summary plots in which standardized expression determined whether higher values increased tumor probability. Feature-ranking stability was quantified as the Jaccard similarity of the top-k SHAP gene sets (k=10, 20, 50) across bootstraps and reported as median and interquartile range.

### Integration of SHAP and differential expression

“Dual-evidence” biomarkers were defined a priori as genes surpassing both statistical dysregulation (FDR<0.05 and |log2FC|≥2) and predictive impact thresholds (top decile of aggregated mean|SHAP| and presence in ≥60% of top-k lists across bootstraps). For visualization, a paired heatmap aligned aggregated mean|SHAP| (z-scaled) with DESeq2 log2FC and FDR overlays. Global concordance between predictive impact and dysregulation was summarized by the Spearman correlation between mean|SHAP| and |log2FC| across expressed genes.

### Statistical analysis, software, and reproducibility

All tests were two-sided with FDR control where applicable. To assess model specificity and guard against leakage, label-permutation experiments (n=100 permutations within 10 randomly chosen bootstraps) were used to verify AUROC collapse toward 0.5 under the null. Analyses were conducted in Python 3.11 (xgboost, scikit-learn, shap, numpy, pandas, matplotlib) and R 4.3 (DESeq2, apeglm, sva, clusterProfiler, org.Hs.eg.db, enrichplot, ggplot2). Random seeds were fixed for bootstrapping and training. All parameters, software versions, and scripts are recorded in a manifest to enable end-to-end re-execution.

## Results

### Cohort and overview

A total of 146 pancreatic tissue RNA-sequencing profiles (74 PDAC, 72 normal) from the TCGA Pan-Cancer Atlas were analyzed using a prespecified pipeline integrating differential gene expression (DGE), pathway enrichment, supervised classification, and model explainability.

### Differential gene expression

Using DESeq2 with false discovery rate (FDR) control (FDR<0.05) and |log₂ fold change| ≥2, tumors showed extensive transcriptomic reprogramming relative to normal pancreas. Prominent overexpressed transcripts included GJB3 (connexin 31), SLC4A11, S100A2, LEMD1, PHLDA2, PAEP, MSLN, and SLC2A1 (GLUT1). Strongly suppressed genes encompassed DEFA6, APOB, RBP2, LCT, MTTP, ASAH2, and multiple solute carriers. These patterns are consistent with loss of exocrine identity, nutrient handling, and epithelial transport in PDAC, alongside induction of epithelial/adhesion and stress-response programs.

### Functional enrichment

Over-representation analyses (GO Biological Process and KEGG) segregated tumor-enriched versus normal-enriched biology. Tumors exhibited activation of epithelial and adhesion programs (keratinocyte/epidermis development, negative regulation of peptidase activity, hemidesmosome assembly), whereas processes characteristic of healthy exocrine tissue—chylomicron assembly, intestinal lipid absorption and cholesterol regulation, carbohydrate/fructose transport, and heme/porphyrin catabolism—were diminished. The aggregate signal indicates epithelial remodeling coupled to extinction of specialized pancreatic metabolic functions.

### Classifier performance

An extreme gradient boosting (XGBoost) classifier trained on normalized expression achieved high discriminative performance across 500 stratified bootstraps: mean area under the ROC curve (AUC) 0.9868 (SD 0.0116), accuracy 0.9763 (SD 0.0136), precision 0.9856 (SD 0.0177), recall 0.9656 (SD 0.0311), and F1 score 0.9751 (SD 0.0140).

Aggregated confusion matrices showed more than 96% correct classification for both classes with infrequent type I/II errors. A parsimony analysis retraining on only the top SHAP-ranked features preserved near-maximal discrimination (AUC ≈0.98).

### Model interpretability and gene prioritization

SHapley Additive exPlanations (SHAP) consistently prioritized GJB3, LINC02086, and TSPAN1 as high-impact contributors, with additional recurrent features including GJB4, HOXC10, SCGB1C1, SLC4A11, and S100A11. SHAP summary plots indicated that higher expression of leading features shifted predictions toward the tumor class, providing directionality and enhancing biological interpretability.

### Integrated “dual-evidence” biomarkers

Intersecting high-impact SHAP features with significantly dysregulated genes identified convergent candidates with both statistical dysregulation and predictive impact. Among these, GJB3 and LINC02086 demonstrated strong mean SHAP influence together with marked overexpression in tumors, supporting their candidacy as diagnostically informative and mechanistically relevant biomarkers. TSPAN1 showed substantial model impact with borderline differential-testing visibility (consistent with thresholding effects), warranting targeted validation.

### Sensitivity and stability analyses

Performance estimates were stable across bootstraps, and SHAP feature rankings showed high overlap among resamples. A reduced top-genes model retained AUC ≈0.98, supporting feasibility of parsimonious panels. Label-permutation checks confirmed collapse of AUC toward 0.5 under the null, arguing against information leakage.

**Figure 1.**
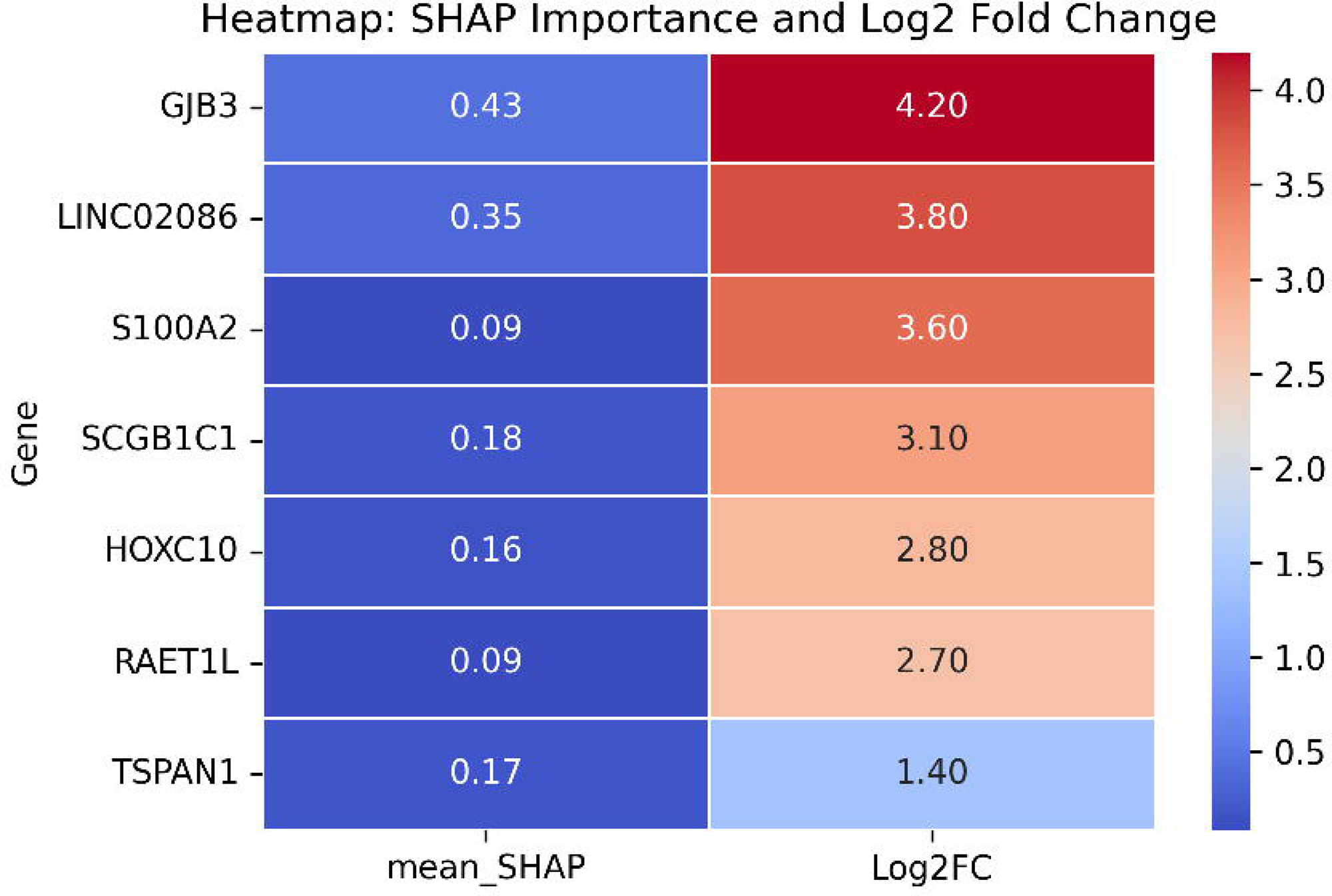
Functional characterization of PDAC transcriptomic alterations. (A) Volcano plot of DESeq2 results using apeglm-shrunk log2FC (FDR<0.05; |log2FC|≥2); red and blue denote up- and down-regulated genes, respectively. (B) Gene–term network for significant GO Biological Processes (q<0.05), with node size reflecting gene count and edge thickness reflecting membership; colors indicate tumor- versus normal-enriched processes after redundancy pruning. (C) Dot plot of GO/KEGG enrichment summarizing gene ratio versus –log10 FDR for up- and down-regulated gene sets.

**Figure 2.**
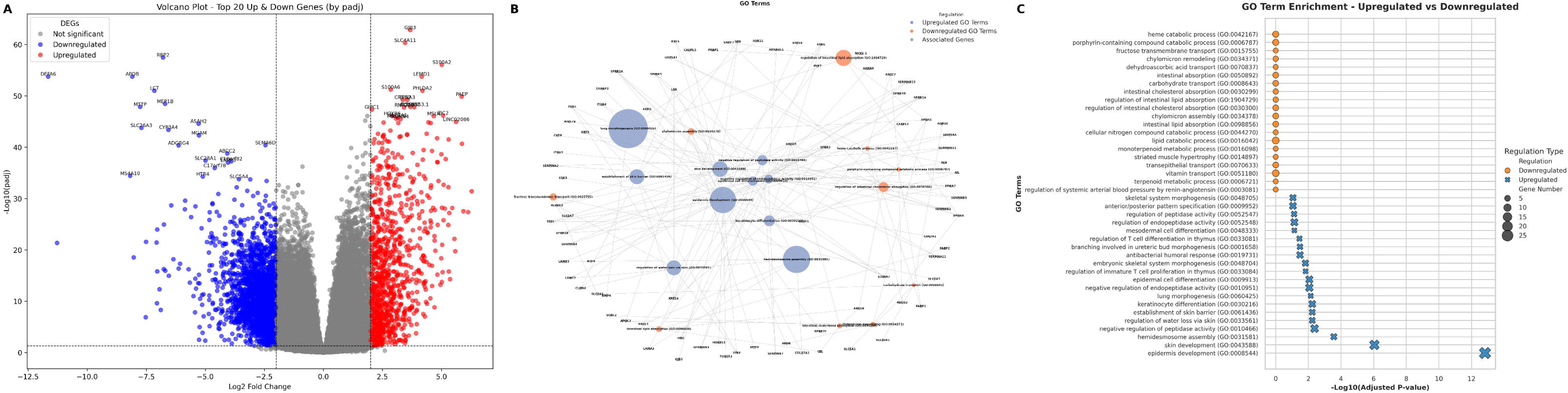
SHAP-based interpretability and classification performance. (A) SHAP summary (beeswarm) for the highest-impact features; points are colored by standardized expression to indicate directionality of effect on tumor probability. (B) Receiver operating characteristic curve for the reduced top-genes model, showing mean AUROC ≈0.98 across bootstraps with a 95% percentile band. (C) Confusion matrix averaged across 500 bootstraps and row-normalized to illustrate error structure. (D) Aggregated mean absolute SHAP values with variability across bootstraps, highlighting frequent high-impact genes (e.g., GJB3, LINC02086, TSPAN1, RAET1L, SLC4A11, HOXC10).

**Figure 3.**
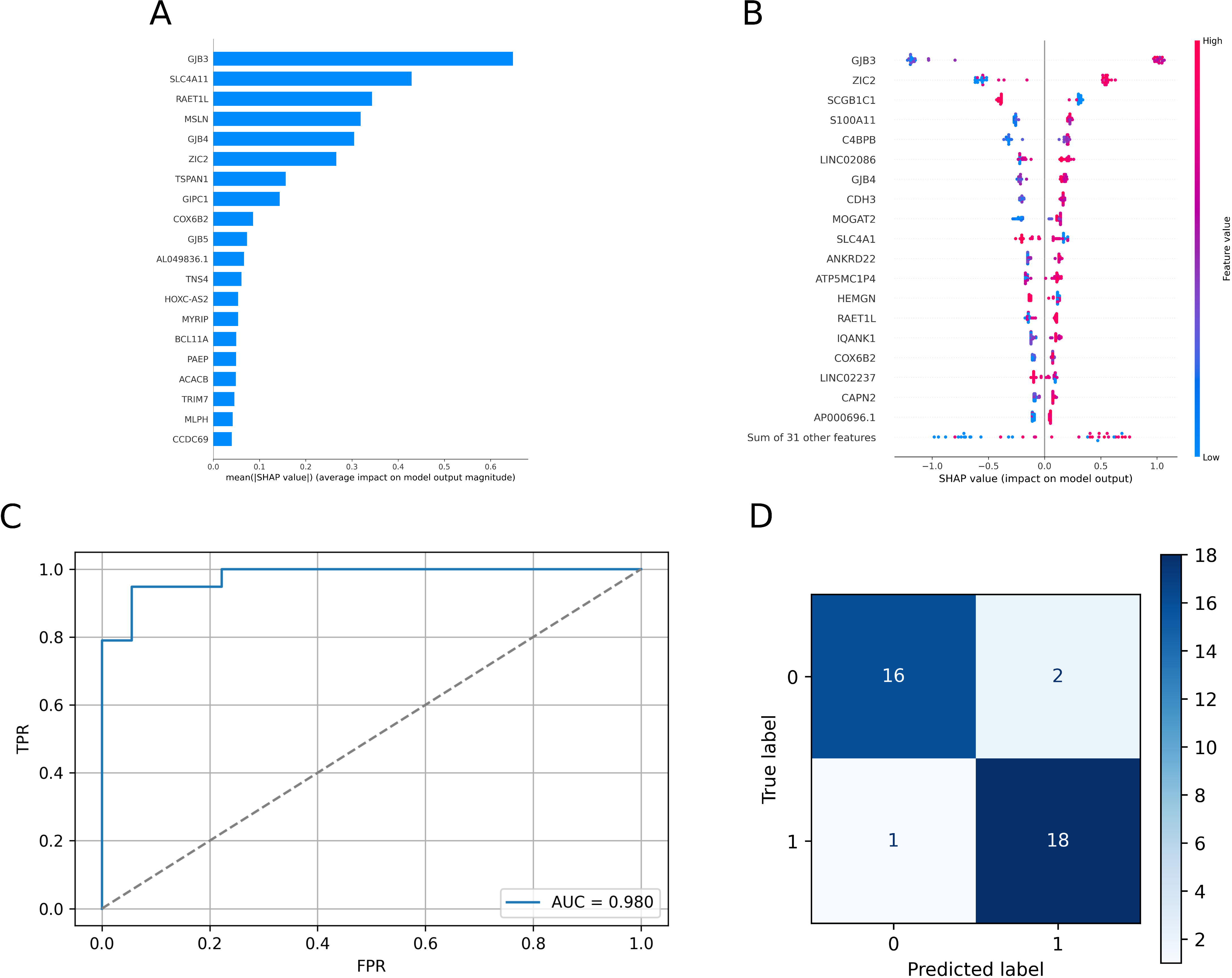
Coupling predictive impact with dysregulation. Paired heatmap aligning, for SHAP-ranked genes, aggregated mean absolute SHAP (z-score) with DESeq2 log2FC and FDR overlay. Genes exceeding both thresholds are highlighted as dual-evidence candidates; GJB3 and LINC02086 display strong predictive contribution and marked overexpression in tumors. Heatmap displaying the mean SHAP values (left column) and corresponding log₂ fold changes (right column) for the top predictive genes identified by the XGBoost model. This integrative visualization highlights genes that exhibit both high model impact and significant differential expression. Notably, GJB3 and LINC02086 demonstrate strong predictive relevance and marked overexpression in tumor tissue, underscoring their potential as dual-significance biomarkers in PDAC.

## Discussion

This study provides an integrative transcriptomic and computational analysis of pancreatic ductal adenocarcinoma (PDAC), combining differential gene expression profiling with interpretable machine learning to identify biologically and diagnostically relevant molecular signatures. The observed dysregulation of genes such as GJB3, S100A2, and SLC2A1, alongside the downregulation of DEFA6, APOB, and RBP2, reflects a profound loss of pancreatic functional identity and the activation of aberrant epithelial differentiation programs. These alterations are consistent with prior evidence indicating exocrine dysfunction, dedifferentiation, and lineage plasticity as hallmarks of PDAC progression [14,15]. Such transcriptional shifts likely underlie the metabolic reprogramming and desmoplastic remodeling that contribute to the tumor’s aggressiveness, metastatic potential, and resistance to therapy [16,17].

Functional enrichment analyses further revealed enrichment of biological processes related to epithelial morphogenesis and keratinization, coupled with suppression of pancreatic-specific metabolic and secretory functions. This transcriptional reprogramming suggests a shift from a differentiated exocrine phenotype toward a progenitor-like, quasi-mesenchymal state, which may foster tumor cell adaptability and treatment evasion [18,19]. Previous studies have shown that loss of pancreatic lineage markers, including PTF1A and NR5A2, correlates with tumor dedifferentiation and poor clinical outcomes [20]. Therefore, the genes identified here not only serve as diagnostic candidates but may also illuminate key molecular mechanisms driving PDAC biology.

The implementation of an XGBoost classifier integrated with SHAP-based interpretability allowed for the robust identification of predictive genes with high diagnostic value. The model achieved an AUC of 0.9868 and a classification accuracy of 98.6%, underscoring the discriminative power of the selected features. Importantly, GJB3, LINC02086, and TSPAN1 emerged as the top contributors to classification performance, with GJB3 and LINC02086 also showing consistent overexpression in tumor tissues. These dual-significance genes—both statistically influential and biologically relevant—represent promising biomarker candidates for early disease detection and mechanistic exploration. The integration of explainable machine learning (XAI) approaches, such as SHAP, enables transparent feature attribution and enhances the interpretability of complex transcriptomic models, bridging the gap between computational predictions and biological insight [21,22].

From a clinical perspective, the identification of reliable biomarkers remains a critical unmet need in PDAC management. Current international guidelines from the NCCN, ESMO, and ASCO advocate a multidisciplinary approach combining surgical resection, systemic chemotherapy, and, in selected cases, radiotherapy or targeted therapy [23-25]. However, treatment decisions—particularly regarding neoadjuvant and adjuvant strategies—are largely based on anatomical resectability and performance status, rather than molecular characteristics. The biomarkers highlighted in this study could refine risk stratification, potentially guiding the selection of patients who might benefit most from intensive perioperative therapy or closer surveillance. For instance, GJB3 overexpression has been linked to enhanced cell–cell communication and metastatic potential, suggesting possible implications for tumor dissemination and chemoresistance [26]. Similarly, LINC02086 and TSPAN1 have been implicated in epithelial-to-mesenchymal transition and may represent new therapeutic targets for pathway inhibition.

Moreover, understanding the transcriptomic reprogramming underlying PDAC could inform the development of novel treatment strategies. The restoration of pancreatic functional programs, or targeted inhibition of aberrantly activated epithelial pathways, might complement existing cytotoxic and targeted therapies. Recent efforts integrating high-throughput sequencing, spatial transcriptomics, and AI-driven modeling have begun to uncover vulnerabilities within the PDAC microenvironment, including stromal–tumor crosstalk and immune evasion mechanisms [27]. These advances underscore the transformative potential of combining omics technologies with explainable machine learning to accelerate biomarker discovery and precision oncology in PDAC.

In conclusion, this study adds to the growing evidence supporting the integration of machine learning into cancer transcriptomics. By combining differential gene expression analysis with SHAP-enhanced feature interpretation, we identified robust, biologically meaningful molecular candidates that may serve as dual-purpose biomarkers—diagnostic and mechanistic—in PDAC. Future validation in independent clinical cohorts and functional studies will be essential to confirm their prognostic and therapeutic relevance. Ultimately, this integrative framework exemplifies a path toward biologically interpretable, AI-guided discovery in one of the most challenging malignancies in modern oncology.

**Limitations** warrant careful consideration. First, this is a single-cohort analysis of publicly available bulk RNA-seq; without external, multi-platform validation, estimates may be optimistic. Second, bulk profiles conflate tumor and stromal compartments, and we did not explicitly adjust for tumor purity or cell-type composition; single-cell and deconvolution analyses will be important to localize signal. Third, although we used nested preprocessing and extensive resampling, cross-sectional design precludes assessment of temporal dynamics, prognosis, or treatment prediction. Fourth, we did not evaluate assay performance on clinical biospecimens (for example, EUS-FNA, FFPE, or liquid biopsy), and platform effects (qPCR versus targeted RNA-seq) may influence performance. Finally, biologic assertions remain inferential; functional studies are needed to establish causality.

**Clinical and public health implications** include the feasibility of translating a parsimonious, interpretable panel into a targeted RNA assay for small specimens, with predefined thresholds informed by SHAP directionality; pairing such an assay with decision-curve analysis and reclassification metrics could quantify clinical utility in perioperative pathways. Integration with circulating analytes (for example, cfRNA) may enable noninvasive screening in high-risk cohorts and longitudinal surveillance.

## Conclusion

Pancreatic ductal adenocarcinoma (PDAC) is characterized by extensive transcriptomic reprogramming, encompassing both the loss of pancreatic-specific functional identity and the activation of aberrant cellular programs. By integrating differential gene expression analysis with interpretable machine learning, we identified a robust molecular signature with high diagnostic potential and biological relevance.

The XGBoost model, complemented by SHAP-based interpretability, demonstrated excellent classification performance and highlighted key predictive genes, including GJB3, LINC02086, and TSPAN1. Notably, several of these genes also exhibited significant dysregulation in tumor tissue, reinforcing their potential as dual-purpose biomarkers. The convergence of statistical significance and model impact positions these candidates as promising targets for further functional validation and potential therapeutic exploration.

Integrating these findings with current standards of care in pancreatic cancer underscores the value of precision medicine approaches. Identified biomarkers, such as GJB3 and LINC02086, could serve as valuable tools to complement existing diagnostic and therapeutic strategies, including surgery, systemic chemotherapy, and radiotherapy [14–18]. Transcriptomic-based patient stratification has the potential to optimize treatment selection, enhancing efficacy while minimizing toxicity in a clinical landscape where therapeutic options are limited and disease progression is rapid [19,20].

Furthermore, the identification of these genes as potential therapeutic targets opens new avenues for drug development. Understanding the molecular underpinnings of functional reprogramming in PDAC is essential to overcome treatment resistance and improve long-term prognosis [5,9]. This study not only validates the utility of combining cancer genomics with artificial intelligence but also provides a framework for translating these discoveries into clinically actionable applications that may ultimately improve patient outcomes.

Overall, our findings emphasize the transformative potential of integrating transcriptomic profiling with explainable machine learning to advance biomarker discovery in PDAC and support the development of precise diagnostic tools for early detection and personalized therapeutic strategies.

## Supplement

### eMethods

#### Cohort, Data Source, and Eligibility

We analyzed bulk RNA-sequencing (RNA-seq) data from the TCGA Pan-Cancer Atlas pancreas cohort comprising 146 human tissues (72 histologically normal pancreas and 74 primary PDAC tumors). The consolidated input file was the unstranded gene-level matrix (balanced_merged_unstranded_pancreas.csv) with gene_id, gene_name, gene biotype, raw counts, and sample-level metadata. Samples were retained if phenotype labels were unambiguous; only primary tumors were included.

#### RNA-seq Processing and Quality Control

Gene-level counts were organized into a sample-by-gene matrix (rows, samples; columns, HGNC symbols) for downstream analysis. Library quality metrics reported in the source repository were reviewed; explicit RIN values were not available. For machine-learning inputs, matrices were transposed to sample-major orientation and standardized as described below.

#### Differential Gene Expression

Differential expression was performed in DESeq2 (via rpy2) using a two-group design (∼ cancer_type). P values were adjusted for multiple testing by the Benjamini–Hochberg false discovery rate (FDR). Significance was prespecified as FDR < 0.05 and |log₂ fold change (log₂FC)| > 2. Shrunken effect sizes (apeglm) were used for visualization and ranking. Results were exported as a fully annotated table and displayed with volcano and MA plots reflecting the stated thresholds.

#### Functional Enrichment

Over-representation analysis (ORA) was conducted with GSEApy on the expressed-gene universe using GO Biological Process 2021 and KEGG Human Pathways 2021. Up- and downregulated DEG lists were analyzed separately, with significance set at adjusted P < 0.05. Enrichment was summarized in tables and depicted with bubble and network plots.

#### Machine-Learning Pipeline

Binary classification (tumor vs normal) used XGBoost trained on variance-stabilized expression matrices. Prior to modeling, features underwent a Quantile Transformer to normal output margins followed by per-gene standardization. Low-variance features were removed using a relative variability threshold of 0.01 to retain informative genes. Model performance was evaluated with 500 stratified bootstrap iterations (75/25 train–test splits per iteration). Within each iteration, accuracy, precision, recall, F1 score, and area under the ROC curve (AUC) were computed and aggregated.

#### Explainability and Stability

Model interpretability relied on SHapley Additive exPlanations (SHAP). SHAP values were computed on the held-out test sets for a defined subset of the initial bootstrap runs, and importance was visualized with summary and beeswarm plots to assess per-gene impact and directionality. Aggregate ROC curves, confusion matrices, and SHAP figures were saved for reporting. Formal list-overlap stability metrics (e.g., Jaccard indices) were not reported in the source analyses.

#### Integrated SHAP × DE Comparison

To nominate “dual-evidence” candidates, predictive impact (mean absolute SHAP values) was integrated with statistical dysregulation (|log₂FC| and FDR). Intersections between SHAP-ranked features and DEG sets were summarized with Venn diagrams and bivariate heatmaps aligning model impact with expression change.

#### Outputs and Reproducibility

Deliverables include the fully annotated DE table, enrichment results, aggregate confusion matrix, mean ROC curve, and SHAP-derived importance visualizations stored in the analysis output directory. Where applicable, CSV exports accompany each analysis step to support end-to-end reproducibility.

### Supplement — eResults

#### Performance Distributions

Across 500 stratified bootstrap iterations (75/25 train–test), the XGBoost classifier achieved mean (SD) accuracy 0.9763 (0.0136), precision 0.9856 (0.0177), recall 0.9656 (0.0311), F1 0.9751 (0.0140), and ROC AUC 0.9868 (0.0116). The aggregated confusion matrix indicated >96% correct classification for both classes with few type I/II errors. A reduced refit using only the top SHAP-ranked genes preserved near-maximal discrimination (AUC ≈0.98).

#### Feature Importance and Explainability

SHAP values computed on held-out test sets (subset of the first five bootstrap runs) identified recurrent high-impact features, notably GJB3, LINC02086, and TSPAN1. Summary/beeswarm plots encoded directionality, showing that higher expression of leading features increased tumor probability.

#### DEG–SHAP Integration (“Dual-Evidence” Candidates)

Intersecting SHAP-prioritized features with statistically significant DEGs yielded a dual-evidence set. GJB3 and LINC02086 displayed both high SHAP impact and marked overexpression, whereas TSPAN1 showed high SHAP impact with comparatively modest log₂ fold change, suggesting non-linear or subgroup-specific effects captured by the model.

#### Functional Signal (Context for Figures)

Differential expression (FDR<0.05; |log₂FC|>2) delineated strong up- and down-regulation consistent with induction of epithelial/adhesion programs and suppression of exocrine transport/metabolic functions. GO/KEGG ORA was run separately for up- and down-regulated sets (adjusted P<0.05) and summarized with dot and network plots.

### Supplement — eTables

**eTable 1. Sample Characteristics and Data Source (TCGA Pan-Cancer Pancreas)**

Columns: Sample_ID; Phenotype (Normal/Tumor); Source (TCGA Pan-Cancer).

*eTable1.merged_tpm_unstranded_pancreas.csv*

**eTable 2. Full Differential Expression Results (DESeq2)** Columns: Gene; baseMean; log2FC (apeglm); SE; Wald_stat; p_value; FDR; Direction (Up/Down). Thresholds: FDR<0.05; |log₂FC|>2.

*eTable2.deseq2_results_annotated.csv*

**eTable 3. GO/KEGG Over-Representation Results (ORA)**

Columns: Collection (GO BP / KEGG); Term_ID; Term_Name; GeneSet_Size; Gene_Ratio; Adjusted_P; Direction (Tumor/Normal); Leading_Genes. Analyzed separately for up- and down-regulated sets.

*eTable3.1.downregulated_GO_Biological_Process_2021_results.csv*

*eTable3.2.upregulated_GO_Biological_Process_2021_results.csv*

**eTable 4. Classification Metrics Across 500 Stratified Bootstraps**

Per-iteration performance of the XGBoost classifier under 500 stratified bootstrap resamples (75/25 train–test splits). For each bootstrap, AUROC, AUPRC, Accuracy, Precision, Recall, F1, Brier score, Specificity, Sensitivity, Threshold at Youden’s J, and class prevalence in the test set were computed and recorded by Bootstrap_ID. Summary rows report the mean (SD) and the 2.5th/97.5th percentiles across bootstraps.

*Abbreviations:* AUROC, area under the receiver operating characteristic curve; AUPRC, area under the precision–recall curve; F1, harmonic mean of Precision and Recall; Brier, mean squared error of probabilistic predictions; Sensitivity (Recall), true-positive rate; Specificity, true-negative rate. *Threshold at Youden’s J* denotes the probability cutoff maximizing (Sensitivity + Specificity − 1). *Class_Prevalence_Test* is the proportion of PDAC samples in each held-out test split.

*eTable4.Classification Metrics Across 500 Bootstraps.csv*

**eTable 5. Dual-Evidence Panel (DEG ∩ SHAP-Top)**

This table lists genes that are both differentially expressed (tumor vs normal) and influential in the classifier. Columns report effect size (log2FC), multiple-testing–adjusted significance (FDR), the average absolute SHAP impact (mean∣SHAP∣, model-scale units), the predominant contribution direction relative to tumor (SHAP_Direction), and a brief functional note.

Columns: Gene; log2FC; FDR; mean(|SHAP|); SHAP_Direction (↑tumor/↓tumor); Brief_Biological_Note.

*eTable 5. Dual-Evidence Panel.SHAPvsLOG2FC.csv*

## Supplement — eFigure Legends

**eFigure 1. Volcano Plot (DESeq2; FDR<0.05; |log₂FC|>2)**

–log₁₀(FDR) vs log₂FC highlighting the top significantly up- and down-regulated genes in PDAC vs normal pancreas.

**eFigure 2. GO/KEGG Enrichment Dot Plot**

Gene ratio vs –log₁₀(adjusted P) for tumor-enriched and normal-enriched terms; dot size encodes set size.

**eFigure 3. GO Biological Process Network (ORA)**

Redundancy-pruned term–gene network; node color encodes direction up & downregulated (Tumor vs Normal).

**eFigure 4. ROC Curve Using SHAP-Top Genes Only**

ROC for the classifier trained exclusively on SHAP-top features, showing AUROC ≈0.98.

**eFigure 5. Aggregated Confusion Matrix (500 Bootstraps)**

Row-normalized confusion matrix averaged across iterations; per-class misclassification rates are low.

**eFigure 6. SHAP Beeswarm (Held-Out Tests)**

Distribution of SHAP values for top features; points colored by standardized expression (red=high, blue=low).

**eFigure 7. Bivariate Heatmap: mean(|SHAP|) and log₂FC**

Paired panels showing SHAP impact (z-scores) and DESeq2 log₂FC with FDR overlay; dual-evidence genes highlighted.

**eFigure 8. Venn Diagram (SHAP-Top ∩ DEGs)**

Overlap between SHAP-ranked features and significant DEGs; intersection genes enumerated in eTable 5.

## Data and Code Availability

De-identified gene-level resources and analytic artifacts generated for this study will be made publicly available upon publication at a persistent repository (DOI to be provided upon acceptance). Released files will include, subject to data-use permissions, gene-level normalized expression matrices; the full differential expression table (deseq2_results_annotated.csv); Gene Ontology and KEGG over-representation results (downregulated_GO_Biological_Process_2021_results.csv) and upregulated_GO_Biological_Process_2021_results.csv); SHAP summaries aggregated across evaluation runs (shap_values_bootstrap_summary.csv) and per-bootstrap classification metrics (Classification Metrics Across 500 Bootstraps.csv). Reproducible code will be deposited alongside the data, comprising R scripts for DESeq2 and clusterProfiler workflows and Python scripts for model training and explainability with XGBoost and SHAP.

## Article Information

### Author Contributions

Concept and design: López-Aladid R, Maciá Escalante S.

Acquisition/curation of data: López-Aladid R.

Technical and computational analysis (all pipelines, statistics, and modeling): López-Aladid R (lead).

Interpretation of data: all authors.

Critical revision of the manuscript for important intellectual content: All authors. Statistical/computational analysis: López-Aladid R.

Administrative/technical/material support: None reported.

Supervision: Maciá Escalante S.

## Data Access, Responsibility, and Analysis

López-Aladid R had full access to all the data in the study and takes responsibility for the integrity of the data and the accuracy of the data analysis. Maciá Escalante S verified analytic outputs and contributed to interpretation.

## Ethical Approval and Patient Consent

This research used de-identified, publicly available RNA-seq data; institutional review board approval and informed consent were not required.

## Data Sharing Statement

The study used de-identified, publicly available RNA-sequencing data from TCGA (pancreas cohort). Derived analytic artifacts—including full differential expression tables, enrichment results, bootstrap metrics, SHAP summaries, and the reduced-panel gene list. Researchers may reproduce the results by using the scripts on the publicly accessible TCGA inputs. Input data were derived from public repositories. Analytical artifacts (code, parameter files, full DE tables, enrichment results, validation metrics, SHAP summaries, and the dual-evidence panel) will be available upon responsbale request. Key files:

1. eTable1.merged_tpm_unstranded_pancreas.csv
2. eTable2.deseq2_results_annotated.csv
3. eTable3.1.downregulated_GO_Biological_Process_2021_results.csv
4. eTable3.2.upregulated_GO_Biological_Process_2021_results.csv
5. etable4.Classification Metrics Across 500 Bootstraps.csv
6. eTable 5. Dual-Evidence Panel.SHAPvsLOG2FC.csv

## Additional Contributions

SMED Clinical Research provided medical writing services support.

## Meeting Presentation

MAP Paris (ESMO), France, 2025, poster format number FPN:8P.

## Acknowledgments

SMED Clinical Research provided medical writing services support.

